# A *KIAA0586* c.1815G>A Founder Allele Associated with Short-Rib Polydactyly Syndrome Type IV in Romani Families

**DOI:** 10.64898/2026.09.29.26363928

**Authors:** Richard Kellermayer, Muhammad Farooq, Judit Bene, Nursel H. Elcioglu, R. Alan Harris, Reka G. Szigeti, Alexandra Teszas, Niels Tommerup, Lars A. Larsen, Bela Melegh, Kinga Hadzsiev

## Abstract

**Background:** Short-rib polydactyly syndrome type IV (Beemer-Langer syndrome) is a lethal skeletal ciliopathy that can result from biallelic pathogenic variants in *KIAA0586* (TALPID3). Earlier clinical observations suggested an unusually high occurrence of this phenotype in Romani families from the Carpathian Basin. Methods: Whole-exome sequencing was performed in affected individuals from five independent families, and the frequency of *KIAA0586* c.1815G>A was assessed in a Hungarian Romani genetic database of 500 individuals. Results: The affected individuals were homozygous for c.1815G>A, a splice-disrupting variant previously shown to cause exon 14 skipping. Four families self-identified as Roma from Hungary and one affected individual was from Turkey. Six heterozygous carriers were identified among 500 individuals in the Hungarian Romani cohort, corresponding to an allele frequency of 0.6%. This was markedly higher than the frequencies cited from ALFA (0.01%) and gnomAD (0.0014%). The Hungarian-Romani and Turkish affected individuals also shared genotypes at informative markers flanking the variant, supporting a shared ancestral haplotype. Conclusions: These observations strengthen evidence that *KIAA0586* c.1815G>A is a founder allele enriched in at least some Romani populations. The data support consideration of ancestry-informed carrier testing in appropriately counseled families. Historical admixture may explain the occurrence of the same haplotype in a Turkish case, but this remains a hypothesis requiring broader population-genomic confirmation.

## Introduction

Short-rib polydactyly syndromes comprise a genetically heterogeneous group of severe skeletal ciliopathies characterized by a constricted thorax, shortened long bones, polydactyly, and variable central nervous system and visceral abnormalities. *KIAA0586* encodes TALPID3, a centrosomal protein required for primary ciliogenesis and Hedgehog signaling [1]. Pathogenic *KIAA0586* variants span a phenotypic continuum that includes Joubert syndrome and lethal short-rib polydactyly phenotypes [2–4].

In 2006, we reported nine individuals with short-rib polydactyly syndrome type IV from seven Romani families in Hungary and proposed a founder allele on clinical grounds [5]. Subsequent work established c.1815G>A (NM_001244189.1; p.Gln605=) as a splice-disrupting *KIAA0586* variant: although synonymous at the protein-coding level, it affects the final nucleotide of exon 14, causes exon skipping, and generates an abnormal transcript predicted to undergo loss of function [2, 6]. The same variant has subsequently been reported in Romani families from Hungary, Romania, Kosovo, and Italy, with variable manifestations extending toward the oralfacial-digital/Joubert spectrum [5, 6].

Founder effects, serial bottlenecks, endogamy, and heterogeneous gene flow have substantially shaped genetic variation among European Romani populations [7–13]. Recent population-genetic work emphasizes marked substructure among Romani groups [7, 8, 12, 13]; therefore, enrichment observed in one cohort should not be generalized to all Romani populations. Here we combine sequencing of affected individuals, a Romani population database, and flanking-marker data to refine the evidence for a *KIAA0586* c.1815G>A founder haplotype and its potential clinical relevance.

## Materials and Methods

Whole-exome sequencing was performed in available affected individuals with a clinical diagnosis of short-rib polydactyly syndrome type IV from five independent families. Variant interpretation focused on *KIAA0586* after identification of the recurrent c.1815G>A allele. Flanking polymorphisms were compared between a Hungarian-Romani affected individual and the Turkish affected individual [15] to assess evidence for a shared local haplotype.

The University of Pécs Romani genetic database, composed of voluntary participants with selfreported Romani ethnicity, was queried for c.1815G>A. Six carriers were identified among 500 individuals. Allele frequency was calculated as 6 variant alleles among 1,000 chromosomes.

The cohort frequency was compared descriptively with frequencies cited from ALFA and gnomAD; the previously reported chi-square comparison was retained as an exploratory population-frequency comparison.

Because the available dataset does not provide genome-wide ancestry inference, dense haplotype phasing, or formal demographic modeling, historical migration scenarios are presented as hypotheses rather than as demonstrated routes of allele transmission.

## Results

All five affected individuals with adequate DNA were homozygous for *KIAA0586* c.1815G>A. Four were from independent Hungarian Romani families and one was the previously described Turkish case. The Hungarian-Romani and Turkish individuals shared the same genotypes across five flanking polymorphisms spanning approximately 36 kb around the variant (Fig. 1C), consistent with a shared local haplotype. These shared homozygous genotypes support, but do not establish, identity by descent; dense phased genotyping would be required to define the founder haplotype precisely.

**Figure 1.**
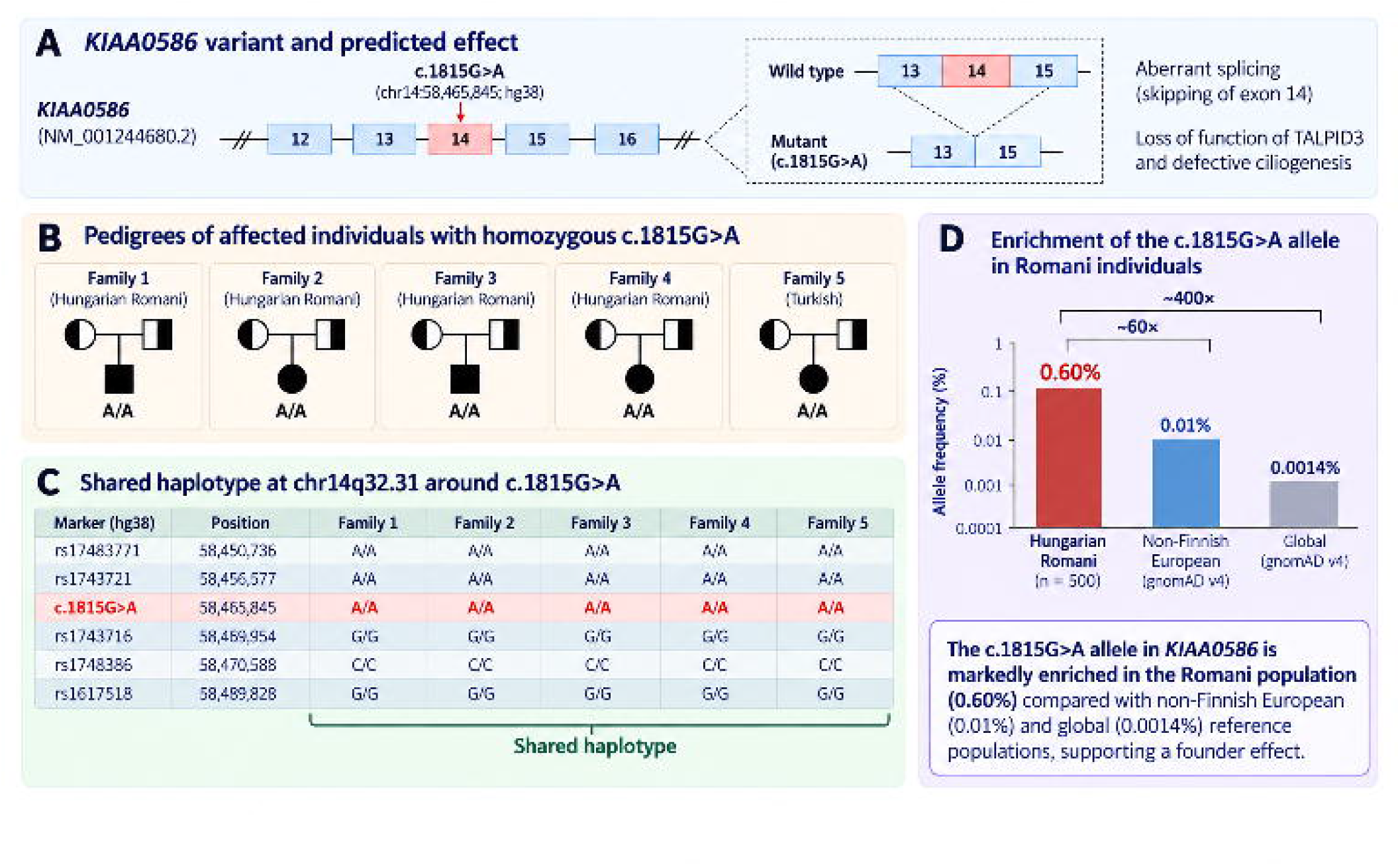
Molecular and population-genetic evidence supporting a recurrent *KIAA0586* c.1815G>A founder allele. (a) Schematic of *KIAA0586* showing c.1815G>A at the terminal nucleotide of exon 14 and the predicted aberrant splicing event with exon 14 skipping, resulting in loss of TALPID3 function and defective ciliogenesis. (b) Pedigree schematics of the five affected families; four are Hungarian Romani families and one is Turkish. Parents are depicted as heterozygous carriers and affected offspring as homozygous for c.1815G>A. (c) Flankingmarker genotypes surrounding c.1815G>A at chr14q32.31, illustrating the local haplotype used to support common ancestry. (d) Allele-frequency comparison showing enrichment of c.1815G>A in the Hungarian Romani cohort (0.60%) relative to the reference frequencies displayed in the figure (0.01% and 0.0014%).

In the Hungarian Romani database, six heterozygous carriers were identified among 500 individuals, corresponding to a carrier frequency of 1.2% and an allele frequency of 0.6%. The comparator frequencies were 0.01% in ALFA and 0.0014% in gnomAD, yielding approximate 60-fold and 400-fold enrichment, respectively (Fig. 1). These comparisons should be interpreted cautiously because reference databases differ in ancestry composition, sampling, and ascertainment.

Prior haplotype analysis of c.1815G>A-positive families from Romania, Hungary, and Kosovo estimated a common ancestor approximately 16 generations (∼480 years) ago. The recurrence of the allele in later reported Romani cases from Italy is compatible with a founder effect followed by population dispersal, although the present study does not establish the precise historical route (Fig. 2) [6, 14].

**Figure 2.**
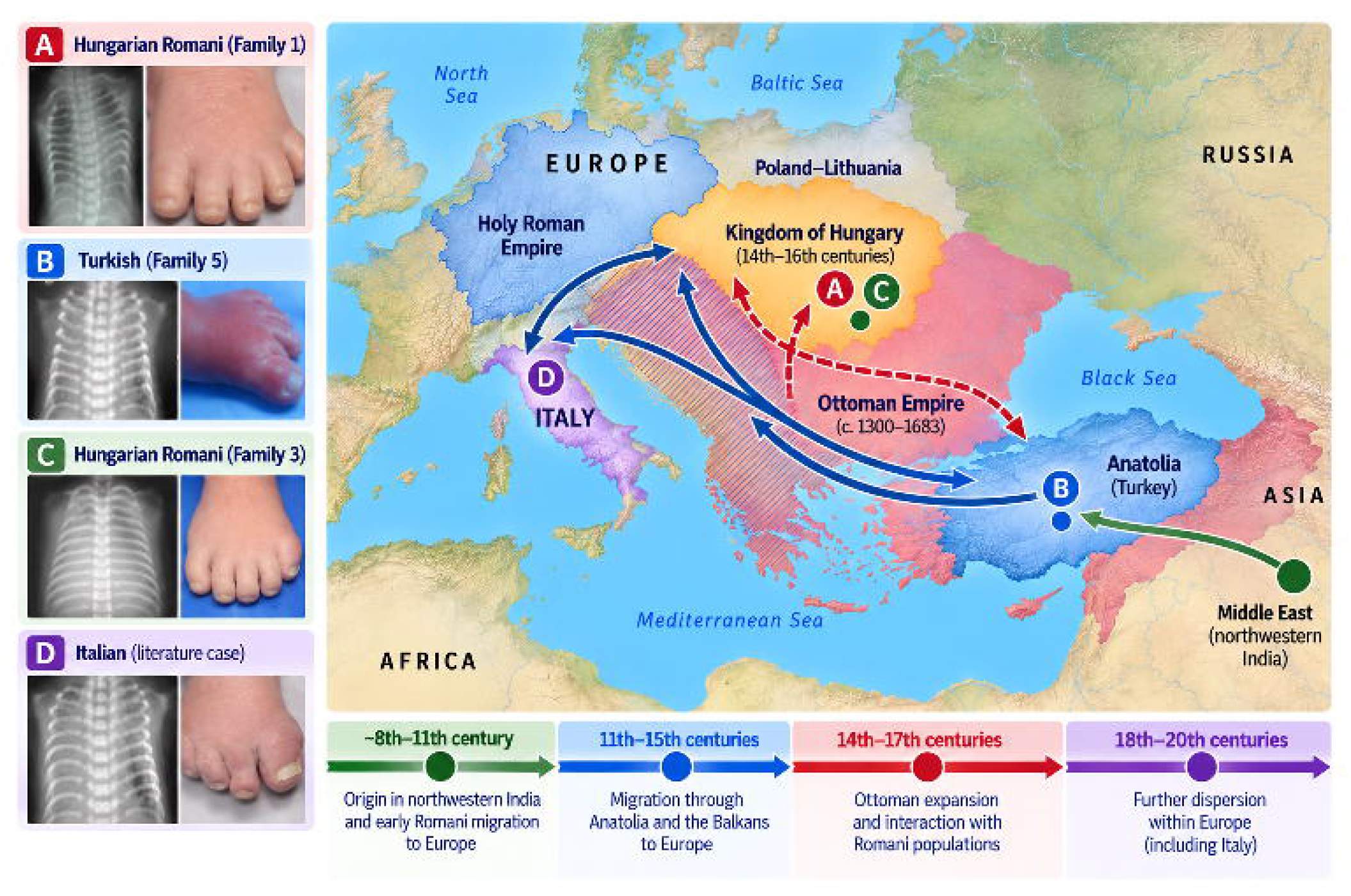
Clinical phenotypes and historically plausible population context of *KIAA0586* c.1815G>A. Clinical panels show representative chest radiographs demonstrating a narrow thorax with short ribs and associated foot abnormalities in (A) Hungarian Romani Family 1, (B) Turkish Family 5, (C) Hungarian Romani Family 3, and (D) an Italian literature case. The map summarizes major historically documented Romani migration through western Eurasia and later dispersal within Europe, together with the geographic origin of the reported cases. The Ottoman-era overlay denotes a historically plausible period of population contact relevant to the Turkish case; it should not be interpreted as demonstrating the direction or timing of transmission of the *KIAA0586* allele. Arrows and century ranges are schematic and represent population-history context rather than a genetically reconstructed migration path for this variant.

## Discussion

The principal finding is the marked enrichment of *KIAA0586* c.1815G>A in the sampled Hungarian Romani population together with recurrence of the same pathogenic allele in geographically separated Romani families and a shared local marker pattern in the Hungarian-Romani and Turkish affected individuals. These observations strengthen the founder-allele interpretation originally suggested by the clinical clustering of short-rib polydactyly syndrome type IV [5].

Enrichment of a recessive pathogenic allele does not require heterozygote advantage. Serial founder events, genetic drift, population bottlenecks, endogamy, and subpopulation structure are well documented in Romani population history and provide a sufficient demographic explanation [7–13]. We therefore regard heterozygote advantage as unproven and do not invoke it as a primary mechanism. The Turkish case is compatible with historical gene flow between populations in southeastern and central Europe, including during and after the Ottoman period [14], but the present genetic data cannot date or localize that admixture event. A specific 16th-century transmission route should therefore be viewed as historically plausible rather than demonstrated (Fig. 2).

Recent genomic studies reinforce the need for this caution. Genome-wide analyses show both a strong founder history and substantial heterogeneity among European Romani groups, with differential local admixture [7–13]. A review of Romani population genetics similarly emphasizes bottlenecks, founder events, substructure, and variable gene flow [13]. In clinical genetics, GeneReviews maintains a resource on founder variants common in Romani populations and notes that founder-variant testing can be useful when a small number of variants account for a substantial proportion of pathogenic alleles in a defined ancestry group [17]. *KIAA0586* c.1815G>A may merit inclusion in such ancestry-informed testing strategies once its frequency and contribution are validated across additional Romani subpopulations.

An intriguing but unresolved observation comes from India. Taori et al. reported a male fetus with short-rib polydactyly syndrome type IV (Beemer-Langer syndrome), diagnosed by prenatal ultrasonography and supported by postnatal radiographic and postmortem examination [16]. Molecular testing was not reported. Given the South Asian ancestry of European Romani populations, it remains unknown whether this phenotypically concordant case carried *KIAA0586* c.1815G>A or represented a genetically distinct form of short-rib polydactyly syndrome. We previously attempted to obtain archival material for retrospective molecular analysis, but no banked tissue from the affected fetus was available. Thus, whether c.1815G>A was already present in the ancestral South Asian population remains an open question.

The study has limitations. The population sample is modest and based on self-reported ethnicity; genome-wide ancestry data were not available. Only a small set of flanking markers was examined, and the Turkish individual cannot establish the direction or timing of gene flow. Comparator frequencies from aggregate reference databases are not ancestry-matched. Finally, the clinical series is enriched for a severe, recognizable phenotype and cannot estimate penetrance across all biallelic genotypes. Broader sequencing of Romani populations, phased haplotype analysis, and ancestry-matched controls would allow formal estimation of founder age, geographic distribution, and carrier frequency.

Despite these limitations, the convergence of recurrent homozygosity in affected families, functional evidence that c.1815G>A disrupts splicing [2, 6], local haplotype sharing, and enrichment in the Hungarian Romani cohort supports a clinically relevant founder effect. For families with relevant ancestry and a history of lethal skeletal ciliopathy, targeted testing for c.1815G>A could be considered as part of genetic counseling, while comprehensive sequencing remains necessary when targeted testing is negative.

## Data Availability

All data produced in the present study are available upon reasonable request to the authors

## Acknowledgments

RK’s work was supported by the Gutsy Kids Fund. RK is a member of the Texas Medical Center Digestive Disease Center, supported by the NIH/NIDDK (P30 DK056338; Center for Gastrointestinal Development, Infection and Injury). Bela Melegh was supported in part by grant NKFIH 138669. The authors acknowledge the late Professor John M. Opitz for his longstanding inspiration in medical genetics.

Generative AI disclosure: OpenAI ChatGPT (GPT-5.6) [18] was used during final manuscript preparation to assist with language editing, organization, reference-formatting checks, and development and refinement of the graphical figures. The authors reviewed the AI-assisted outputs against the source literature and underlying study data, corrected them where necessary, and take full responsibility for the accuracy, originality, interpretation, and final content of the manuscript and figures. Generative AI was not used to generate or alter the clinical radiographs or patient photographs.

## Statement of Ethics

The Hungarian patient-, and the University of Pécs Romani database studies were performed under National Research, Development and Innovation Office grant #NKFIH 138669 approved by the Scientific and Research Ethics Committee of the Medical Research Council of Hungary. The Turkish patient studies we performed under protocol “Large-scale genomic study of genetic skeletal system disorders” 2017/0284, approved by the Medeniyet University Institutional Review Board.

## Author Contributions

Author contribution details should be completed by the authors before submission in accordance with Karger authorship requirements.

## Data Availability Statement

The data supporting the findings of this study are not publicly available because they include potentially identifiable human genetic information. Further inquiries may be directed to the corresponding author, subject to applicable ethical and institutional restrictions.

## References

1 Bangs F, Antonio N, Thongnuek P, et al. Generation of mice with functional inactivation of talpid3, a gene first identified in chicken. Development. 2011;138:3261–72.

2 Alby C, Piquand K, Huber C, et al. Mutations in KIAA0586 cause lethal ciliopathies ranging from a hydrolethalus phenotype to short-rib polydactyly syndrome. Am J Hum Genet. 2015;97:311–8.

3 Bachmann-Gagescu R, Phelps IG, Dempsey JC, et al. KIAA0586 is mutated in Joubert syndrome. eLife. 2015;4:e06602.

4 Poretti A, et al. Recurrent, founder and hypomorphic variants contribute to the genetic landscape of Joubert syndrome. J Med Genet. 2023;60:885–95.

5 Kovacs N, Sarkany I, Mohay G, et al. High incidence of short rib-polydactyly syndrome type IV in a Hungarian Roma subpopulation. Am J Med Genet A. 2006;140:2816–8.

6 Cocciadiferro D, Agolini E, Digilio MC, et al. The splice c.1815G>A variant in KIAA0586 results in a phenotype bridging short-rib-polydactyly and oral-facial-digital syndrome: a case report and literature review. Medicine (Baltimore). 2020;99:e19169.

7 Bianco E, Laval G, Font-Porterias N, et al. Recent common origin, reduced population size, and marked admixture have shaped European Roma genomes. Mol Biol Evol. 2020;37:3175–87.

8 Font-Porterias N, Arauna LR, Poveda A, et al. European Roma groups show complex West Eurasian admixture footprints and a common South Asian genetic origin. PLoS Genet. 2019;15:e1008417.

9 Martínez-Cruz B, Mendizabal I, Harmant C, et al. Origins, admixture and founder lineages in European Roma. Eur J Hum Genet. 2016;24:937–43.

10 Mendizabal I, Lao O, Marigorta UM, et al. Reconstructing the population history of European Romani from genome-wide data. Curr Biol. 2012;22:2342–9.

11 Mendizabal I, Valente C, Gusmão A, et al. Reconstructing Roma history from genome-wide data. PLoS One. 2013;8:e58633.

12 Antinucci M, Comas D, Calafell F. Population history modulates the fitness effects of copy number variation in the Roma. Hum Genet. 2023;142:1327–43.

13 López M, et al. Population genetics of the European Roma—a review. Genes (Basel). 2022;13:2068.

14 Banfai Z, Melegh BI, Sumegi K, et al. Revealing the genetic impact of the Ottoman occupation on ethnic groups of East-Central Europe and on the Roma population of the area. Front Genet. 2019;10:558.

15 Elçioğlu N, Karatekin G, Sezgin B, Nuhoğlu A, Cenani A. Short rib-polydactyly syndrome in twins: Beemer-Langer type with polydactyly. Clin Genet. 1996;50:159–163. doi:10.1111/j.1399-0004.1996.tb02373.x.

16 Taori KB, Sharbidre KG, Krishnan V, Kundargi N, Kulkarni BR, Satkar V, et al. Diagnosis of short rib polydactyly syndrome type IV (Beemer-Langer syndrome) with cystic hygroma: a case report. J Clin Ultrasound. 2009;37:406–409. doi:10.1002/jcu.20603.

17 Wallace SE. Resources for Genetics Professionals—Genetic Disorders Associated with Founder Variants Common in the Romani Population. GeneReviews®. Seattle: University of Washington; 2022. Updated online.

18 OpenAI. ChatGPT (GPT-5.6) [large language model]. San Francisco (CA): OpenAI; 2026. Available from: https://chatgpt.com/.

